# Neutralization of VOCs including Delta one year post COVID-19 or vaccine

**DOI:** 10.1101/2021.08.12.21261951

**Authors:** Sebastian Havervall, Ulrika Marking, Max Gordon, Henry Ng, Nina Greilert-Norin, Sarah Lindbo, Kim Blom, Peter Nilsson, Mia Phillipson, Jonas Klingström, Sara Mangsbo, Mikael Åberg, Sophia Hober, Charlotte Thålin

## Abstract

**Background:** SARS-CoV-2 variants, such as Alpha, Beta, Gamma and Delta, are raising concern about the efficiency of neutralizing antibodies (NAb) induced by wild-type infection or vaccines based on the wild-type spike.

**Methods:** We determined IgG and NAb against SARS-CoV-2 variants one year following mild wild-type infection (n=104) and two-dose regimens with BNT162b2 (BNT/BNT) (n=67), ChAdOx1 (ChAd/ChAd) (n=82), or heterologous ChAdOx1 followed by BNT162b2 (ChAd/BNT) (n=116).

**Findings:** Wild type spike IgG and NAb remained detectable in 80% (83/104) of unvaccinated participants one year post mild infection. The neutralizing capacity was similar against wild type (reference), Alpha (0.95 (0.92-0.98) and Delta 1.03 (0.95-1.11) but significantly reduced against Beta (0.54 (0.48-0.60)) and Gamma 0.51 (0.44-0.61). Similarly, BNT/BNT and ChAd/ChAd elicited sustained capacity against Alpha and Delta (1.01 (0.78-1.31) and 0.85 (0.64-1.14)) and (0.96 (0.84-1.09) and 0.82 (0.61-1.10) respectively), with reduced capacity against Beta (0.67 (0.50-0.88) and 0.53 (0.40-0.71)) and Gamma (0.12 (0.06-0.27) and 0.54 (0.37-0.80)). A similar trend was found following ChAd/BNT (0.74 (0.66-0.83) and 0.70 (0.50-0.97) against Alpha and Delta and 0.29 (0.20-0.42) and 0.13 (0.08-0.20) against Beta and Gamma).

**Interpretation:** Persistent neutralization of the wide-spread Alpha and Delta variants one year after wild-type infection may aid vaccine policy makers in low-resource settings when prioritizing vaccine supply. The reduced capacity of neutralizing Beta and Gamma strains, but not the Alpha and Delta strains following both infection and three different vaccine regimens argues for caution against Beta and Gamma-exclusive mutations in the efforts to optimize next generation SARS-CoV-2 vaccines.

**Funding:** A full list of funding bodies that contributed to this study can be found in the Acknowledgements section

## Introduction

Increasing data support that neutralizing antibodies following SARS-CoV-2 infection provide a durable protection against reinfection (1-7), and vaccines have been shown effective in constraining hospitalization and deaths associated with SARS-CoV-2 infection (8-11). However, emerging SARS-CoV-2 variants are raising concerns. Viral variants of concern (VOC) with increased transmissibility, such as B.1.1.7 (Alpha), B.1.351 (Beta), P.1 (Gamma) and B.1.617.2 (Delta), entail spike protein mutations conferring a fitness advantage affecting its biology, infectivity, transmissibility, and antigenicity (Figure 1). Spike alterations may enable immune escape, specially voiced when it comes to the B.1.351 (Beta) and P1 (Gamma) VOC. Both display three mutations in the RBD region and particularly related to the E484K mutation, which has been shown to reduce antibody binding and neutralization (12). Assessing the efficiency of humoral immunity following wild-type infection and current vaccines against these globally spreading VOCs is important, both for understanding SARS-CoV-2 virulence and for the optimization of next generation SARS-CoV-2 vaccines (13-21). Using a high-throughput spike-specific IgG and pseudoneutralization assay, we determined the SARS-CoV-2 IgG antibody levels and neutralization capacity against the wild-type virus and VOCs one year following SARS-CoV-2 wild-type infection or following three different vaccine regimens employing the BNT162b2 (Comirnaty, Pfizer-BioNTech) and ChAdOx1 nCoV-19 (Vaxzevria, AstraZeneca) vaccines.

**Figure 1.**
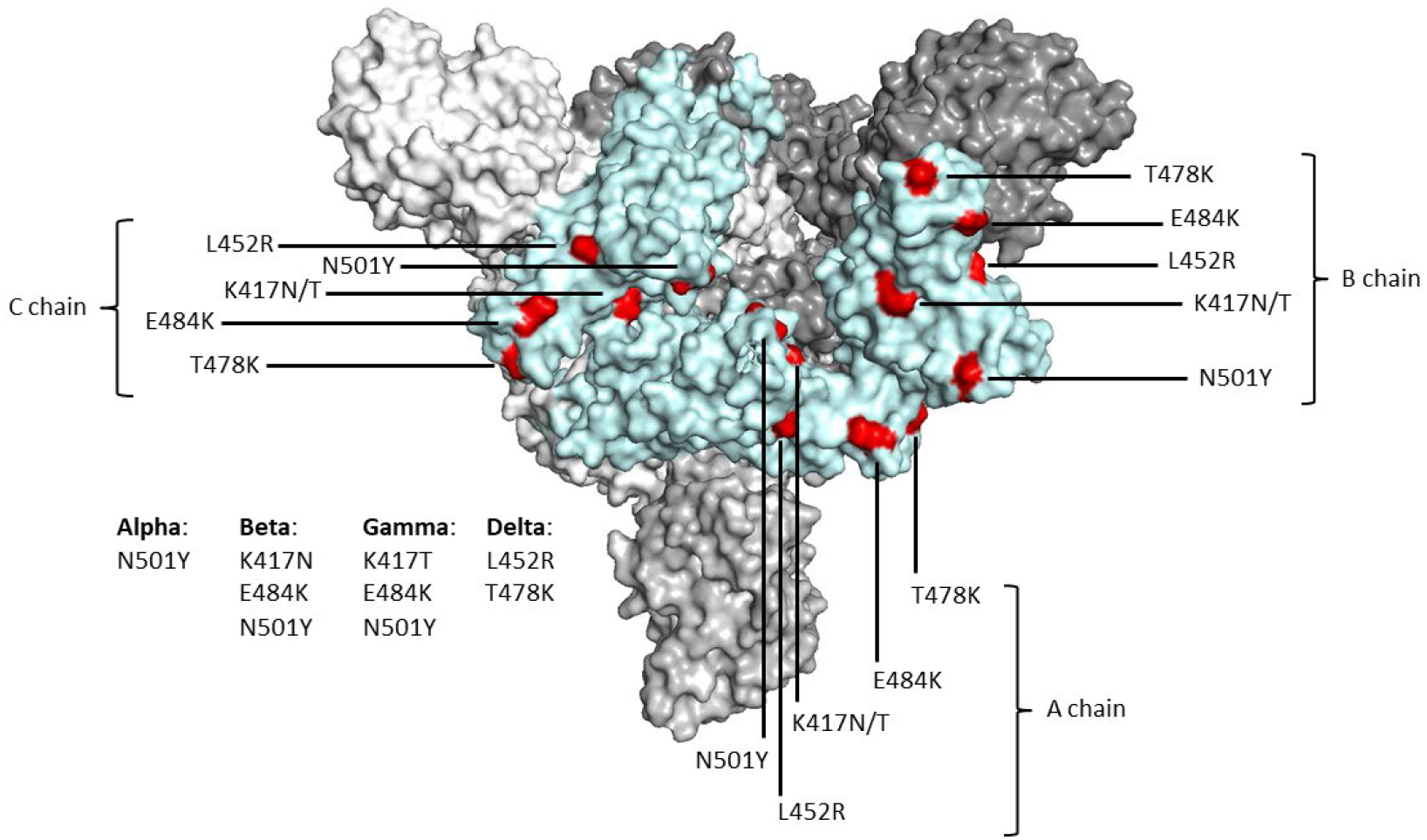
A top view of the structure of a furin-cleaved spike protein of SARS-CoV-2 is shown (6ZGG) with the receptor binding domain (RBD) of the three chains colored in light blue. Positions of RBD that are mutated in the four most common VOCs are colored in red. RBD of the B chain is shown in the erected state (29). The different mutations included in the four VOCs are listed in the figure.

## Methods

The COMMUNITY (COVID-19 Biomarker and Immunity) study (22, 23) investigates immune responses after COVID-19 infection and vaccination. Between April 15 and May 8, 2020, 2149 healthcare workers (HCW) at Danderyd Hospital, Stockholm, Sweden were included. All study participants are invited for repeated blood samplings every four months. Serology was performed using two assays; a multiplex antigen bead assay (FlexMap3D, Luminex Corp) (24) measuring spike-specific IgG and the V-PLEX SARS-CoV-2 Panel 13 (Meso Scale Diagnostics, USA) measuring spike-specific IgG and their neutralizing capacity (pseudo-neutralizing spike-ACE2 assay) against SARS-CoV-2 wild-type and variants B.1.1.7 (Alpha), B.1.351 (Beta), P.1 (Gamma), and B.1.617.2 (Delta). Clinical sensitivity, specificity, and cut-off values for the wild-type strain for both MSD assays (K15463U (IgG), K15466U (ACE2)) were established by ROC analysis of 200 commercially sourced pre-2109 healthy adults and 214 PCR-confirmed COVID-19 patients by the manufacturer. Values for each assay can be obtained from kit insert Appendix G found at the manufacturer’s homepage http://www.mesoscale.com.

This sub study included one group with all study participants that were confirmed SARS-CoV-2 Spike IgG positive by both assays at study inclusion, who attended all follow-ups and who were not yet vaccinated by the 12-month follow-up. All participants reported mild COVID-19 symptoms, and none were hospitalized. Blood samples were also obtained from three groups of vaccinated individuals: all participants who received two doses of BNT162b2 vaccine (BNT × 2; median dose interval 21 days (IQR 21-21), two doses of ChAdOx1 nCoV-19 vaccine (ChAd × 2; median dose interval 81.5 days (IQR 77-84)) or a prime dose of ChAdOx1 nCoV-19 vaccine and booster dose with BNT162b2 vaccine (ChAd + BNT; median dose interval 90 days (IQR 86-94)). Blood samples were collected 30-55 days following the BNT × 2 regimens, and 14-22 days following regimens with ChAd priming. The study was approved by the Swedish Ethical Review Authority (dnr 2020-01653), and written informed consent was obtained from all study participants.

### Statistical analysis

We used mixed effect modelling with subject id as random effect where the outcome during modelling was converted using the log2-function. Interaction between strain and timespans was tested in two subsequent steps, baseline vs any other follow-up, and within the followups. The models were compared using ANOVA. Analyses were performed in R version 4.1.0 with nlme-package (v. 3.1.152). The estimates were compared using the contrast-package (v. 0.22). Each model was adjusted for age and sex. Correlation analysis was performed using Spearman.

#### Role of the funding source

The funders had no role in the design and conduct of the study; collection, management, analysis, and interpretation of the data; preparation, review, or approval of the manuscript; and decision to submit the manuscript for publication.

## Results

A total of 104 study participants who were confirmed spike IgG positive at study inclusion and attended all four-month follow-ups remained unvaccinated at the 12-month follow-up (median age 42 (IQR 31.75-51.25), 88% female). In line with prior reports (3, 4, 25) the levels of both spike binding IgG and neutralizing antibodies (NAb) against the SARS-CoV-2 wild-type strain declined over the first few months, but thereafter remained stable, (Fig 2 A-B), with similar trends for all SARS-CoV-2 VOCs (Table S1). High spike IgG and NAb levels at baseline correlated with high levels 12 months post infection against the wild-type virus and all four VOCs (all r > 0.5 and maximum p < 0.0001). Wild-type spike IgG remained detectable in 91% (95/104), 84% (87/104) and 80% (83/104) of the participants at the 4-, 8- and 12-month follow-up, respectively, with corresponding Wild-type NAb being 96% (100/104), 93% (97/104) and 91% (95/104).

**Figure 2.**
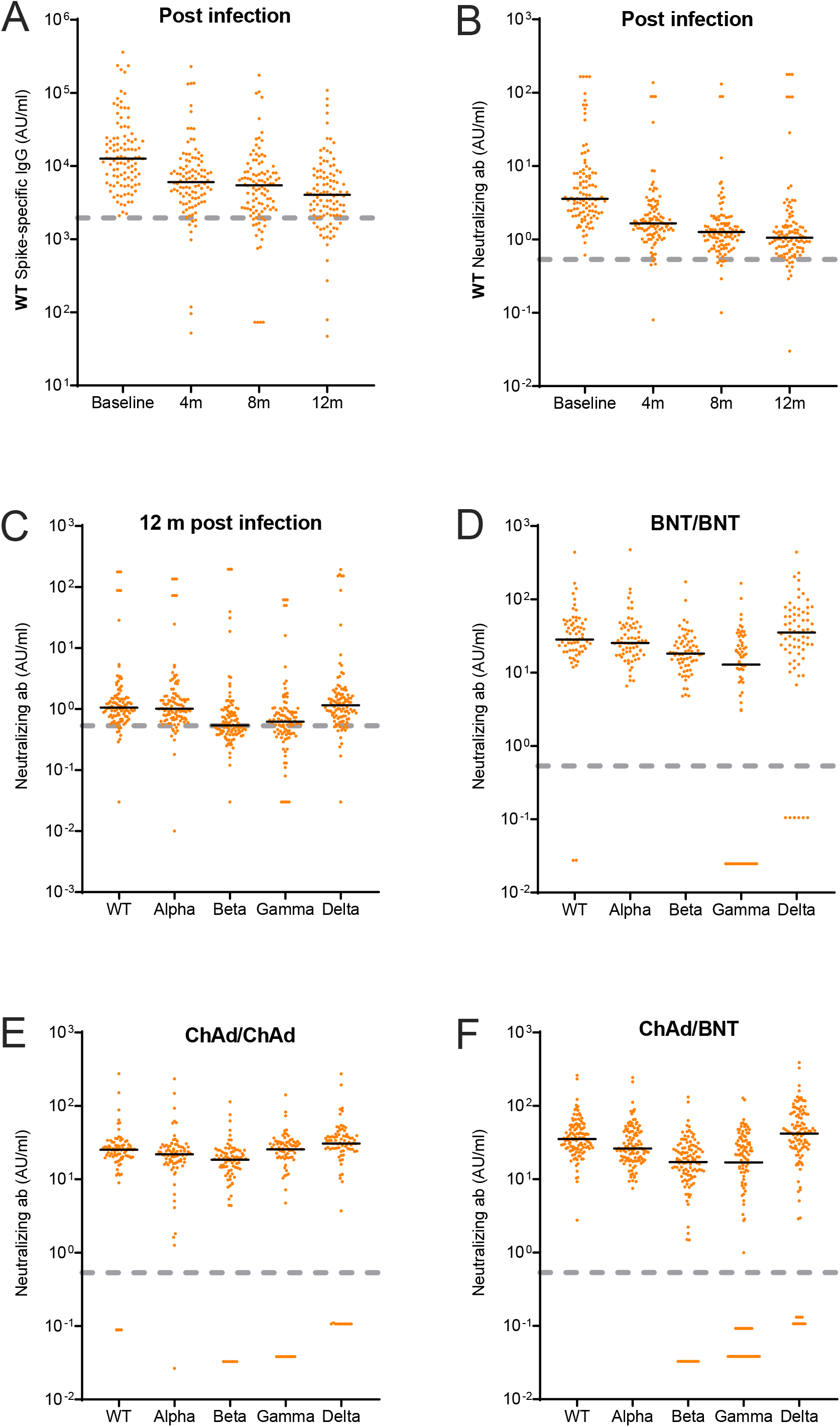
**A-B**. Wild type (WT) spike-specific IgG and neutralizing antibodies (AU/ml) after SARS-CoV-2 WT infection at baseline and at the 4-, 8-, and 12-months follow-up visit. **C-F**. Neutralizing antibodies (AU/ml) for wild type (WT) and B.1.1.7 (Alpha), B.1.351 (Beta), P.1 (Gamma) and B.1.617.2 (Delta) after **C**: 12 months infection, **D**: two doses BNT162b2 vaccine, **E**: two doses ChAdOx1 nCoV-19, **F**: heterologous vaccine regimen with ChAdOx1 nCoV-19 and BNT162b2. Black lines represent medians. Dashed grey lines represent cut-off values for wild type as determined by the manufacturer: IgG = 1960, neutralization = 0.533. WT; Wild type, BNT; BNT162b2 vaccine, ChAd; ChAdOx1 nCoV-19; m; months, ab; antibodies

Differences in neutralizing capacity against the SARS-CoV-2 wild-type and VOCs using samples taken 12 months post infection was determined by quantile regression adjusting for age and sex. With the neutralizing capacity against the SARS-CoV-2 wild-type strain as the reference, the capacity of the antibodies induced by the natural infection of the wild-type virus was only slightly reduced against the B.1.1.7 (Alpha) and similar against the B.1.617.2 (Delta) strain. There was, however, a notable and reduction against the B.1.351 (Beta) and P.1 (Gamma) strains (Figure 2C, 3 and Table S2).

**Figure 3.**
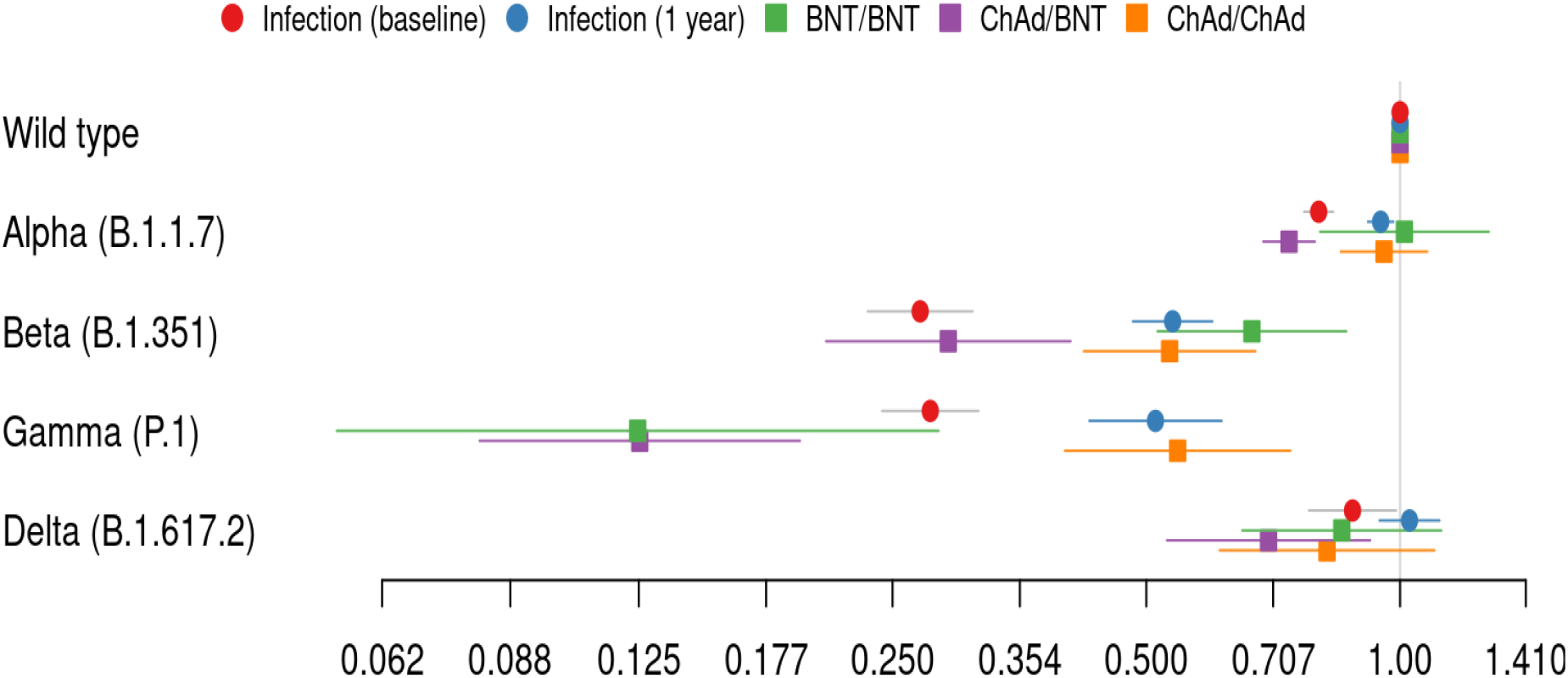
Forest plot of regression output for strain type neutralization. Estimates are multiplicative, i.e. 0.5 corresponds to a 50% reduction in antibody response in relation to the wild-type strain. The model is a mixed effect quantile regression for the median. BNT; BNT162b2 vaccine, ChAd; ChAdOx1 nCoV-19

We next investigated differences in neutralizing capacity against the SARS-CoV-2 wild-type and VOCs following vaccination with either two doses BNT162b2 vaccine (n=67, median age 52 (IQR 41-59), 84% female), two doses ChAdOx1 nCoV-19 vaccine (n=82, median age 51 (IQR 42-57), 88% female) or a prime dose of ChAdOx1 nCoV-19 vaccine and booster dose with BNT162b2 vaccine (n=116, median age 50 (IQR 40-56), 92% female). Similar to the responses observed 12 months post infection, the neutralizing capacity following the BNT/BNT and ChAd/ChAd vaccine regimens were similar against the B.1.1.7 (Alpha), B.1.617.2 (Delta) and wild-type strains. However, there was a reduction against the B.1.351 (Beta) and P.1 (Gamma) strains (Figure 2 D-E, 3 and Table S2). The ChAd/BNT vaccine regimen elicited antibodies capable of similar neutralization against the B.1.617.2 (Delta) and the wild-type strain, but with slightly lower neutralizing capacity against the B.1.1.7 (Alpha) strain. NAb titers against the B.1.351 (Beta) and P.1 (Gamma) strains were, as with the two other vaccine regimens, significantly reduced (Figure 2F and 3 and Table S2).

## Discussion

Our results demonstrate initial reductions but thereafter stable SARS-CoV-2 IgG levels over 12 months following mild SARS-CoV-2 infection. Circulating and readily detectable antibodies are crucial in the assessment and diagnosis of prior infection both on an individual and population-based level. We furthermore show that these antibodies display neutralizing abilities against the B.1.1.7 (Alpha) and B.1.617.2 (Delta) strains similar to those against the wild-type strain, and that this capacity is maintained over time, implying a robust immunity also against these now globally spreading strains. Notably, neutralizing capacity was, however, significantly reduced against the B.1.351 (Beta) and P.1 (Gamma) strains.

The two vaccines used in the studied regimens are both based on the wild-type SARS-CoV-2 spike protein. We found that these three vaccine regimens elicited a similar response as infection with the wild-type strain. Although the virulence of a certain VOC as well as the level of evoked immune protection is dependent on a multitude of features, it is highly influenced by the interaction between the viral spike protein and the host ACE2 receptor. A VOC with higher affinity to ACE2 potentially needs fewer virions to be infective. Further, escape mutants able to evade the immune system might appear as the immunity develops globally. To date, there is published data showing a higher affinity to ACE2 for two (B.1.1.7 (Alpha) and B.1.351 (Beta)) of the four VOCs (26), while data regarding the other two VOCs (P.1 (Gamma), and B.1.617.2 (Delta)) is still missing. Our data indicate no loss of neutralisation of lineage B.1.1.7 (Alpha), suggesting that the higher transmissibility depends on viral characteristics such as the reported higher affinity gained by the N501Y. The additional mutations in positions 417 and 484 are affecting the neutralizing possibility where a threonine in position 417 (in P.1 (Gamma)) appears to give a negative effect for the BNT vaccine (Figure 1 and 3). The higher transmissibility of lineage B.1.617.2 (Delta) does not seem to be based on a lower antibody protection due to the two mutations in RBD alone. However, a higher rate of proliferation has been reported (27) for this VOC. The major concern regarding this VOC does not seem to stem from immune escape but could instead be due to other improved fitness characteristics.

So far, the E484 mutation has been reported to affect antibody binding and neutralization (28) and as such, both B.1.351 (Beta) and P.1 (Gamma) would be at risk of escaping immune control induced by the wild-type strain or current vaccines, which is corroborated by our IgG and Nab results.

Limitations of this study include the observational and single-center design and the large portion of women. Strengths include the long follow-up and different vaccine regimes within the same study protocol along with the baseline sample opportunity against the wild-type and VOC strains.

Our findings of long-lasting neutralization of the widely spreading B.1.1.7 (Alpha) and B.1.617.2 (Delta) variants one year after wild type infection may aid vaccine policy makers in low-resource settings when prioritizing limited vaccine supply. Furthermore, the globally reduced capacity of neutralizing B.1.351 (Beta) and P.1 (Gamma) strains, but not the B.1.1.7 (Alpha) and B.1.617.2 (Delta) strains following both infection and three different vaccine regimens argues for caution against B.1.351 (Beta) and P.1 (Gamma)-exclusive spike protein mutations in the efforts to optimize next generation SARS-CoV-2 vaccines.

## Data Availability

The anonymized datasets generated during and/or analyzed during the current study are available from the corresponding author on reasonable request.

## Acknowledgements

This work was funded by Jonas & Christina af Jochnick foundation; Lundblad family foundation; Region Stockholm; Knut and Alice Wallenberg foundation; Science for Life Laboratory (SciLifeLab); Erling-Persson family foundation; CIMED and the Swedish Research Council.

## Conflict of Interest

SoH has participated on Astra Zeneca COVID-19 SCG Virtual Advisory Board. Otherwise, the authors declare no competing interests.

## Author contribution

SeH, UM, and CT had full access to all the data in the study and takes responsibility for the integrity of the data and the accuracy of the data analysis.

SeH, UM, SoH, and CT designed the study, and drafted the manuscript. MG made the statistical analyses. NG, HN, MÅ conducted the analysis and the administrative and material support. SeH, UM, SL, KB, PN, SM, JK, MP, MÅ, SoH, PN, CT interpreted the data, and all authors critically revised the manuscript. SoH, PN, CT obtained funding. MP, MÅ, SoH, CT supervised. All authors had final responsibility for the decision to submit for publication.

## Supplementary data

**Table S1.**

|  | Baseline |  | 4m |  | 8m |  | 12m |  |
| --- | --- | --- | --- | --- | --- | --- | --- | --- |
|  | Median | IQR | Median | IQR | Median | IQR | Median | IQR |
| IgG (kAU/ml) |  |  |  |  |  |  |  |  |
| Wild type | 12.6 | (6.3-23.3) | 6.0 | (3.6-10.0) | 5.5 | (2.6-8.3) | 4.0 | (2.1-7.6) |
| Alpha (B.1.1.7) | 9.9 | (5.4-17.5) | 5.9 | (3.3-9.9) | 4.9 | (2.6-8.1) | 4.0 | (2.2-7.2) |
| Beta (B.1.351) | 5.4 | (3.2-11.2) | 3.7 | (2.2-6.4) | 3.8 | (1.9-6.5) | 2.8 | (1.6-5.4) |
| Delta (B.1.617.2) | 4.9 | (2.8-12.5) | 3.5 | (2.1-6.1) | 3.5 | (1.8-5.8) | 2.6 | (1.5-5.1) |
| Gamma (P.1) | 6.7 | (3.1-14.0) | 3.5 | (2.1-6.4) | 3.7 | (1.7-6.3) | 2.6 | (1.3-5.0) |
| Neutralizing ab (AU/ml) |  |  |  |  |  |  |  |  |
| Wild type | 3.6 | (2.3-7.9) | 1.7 | (1.2-2.6) | 1.3 | (0.9-1.7) | 1.1 | (0.8-1.5) |
| Alpha (B.1.1.7) | 2.8 | (2.0-6.1) | 1.5 | (1.1-2.5) | 1.2 | (0.9-1.8) | 1.0 | (0.7-1.6) |
| Beta (B.1.351) | 1.1 | (0.8-2.2) | 0.7 | (0.5-1.1) | 0.6 | (0.4-0.9) | 0.5 | (0.4-0.9) |
| Delta (B.1.617.2) | 3.7 | (2.1-7.6) | 1.6 | (0.9-2.3) | 1.1 | (0.6-1.7) | 1.2 | (0.7-1.7) |
| Gamma (P.1) | 1.2 | (0.8-2.4) | 0.8 | (0.4-1.3) | 0.5 | (0.1-0.9) | 0.6 | (0.4-0.9) |
| Median and interquartile range for antibody response for SARS-CoV-2 variants (n=104) |  |  |  |  |  |  |  |  |

**Table S2.**
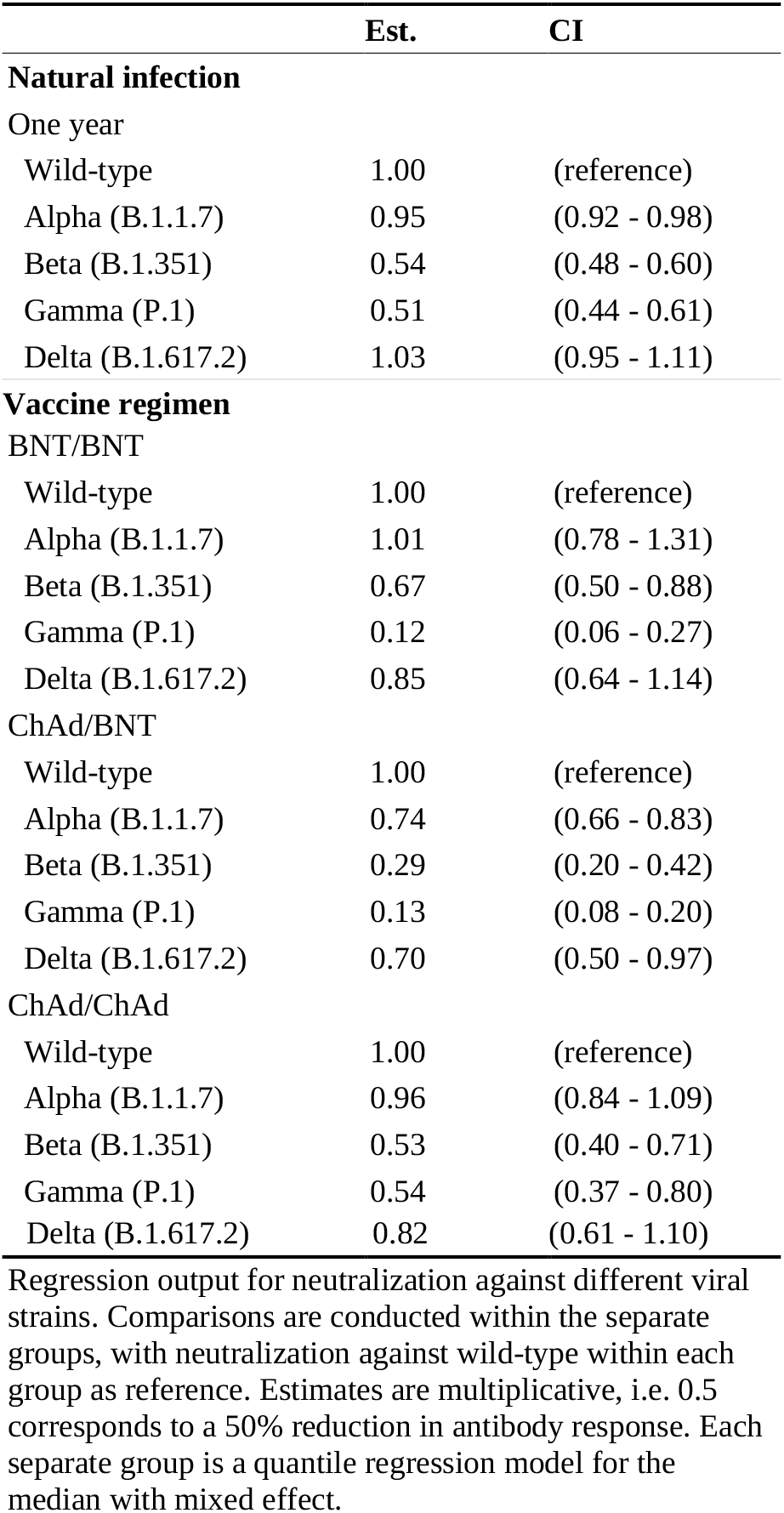

|  | Est. | CI |
| --- | --- | --- |
| **Natural infection** |  |  |
| One year |  |  |
| Wild-type | 1.00 | (reference) |
| Alpha (B.1.1.7) | 0.95 | (0.92 - 0.98) |
| Beta (B.1.351) | 0.54 | (0.48 - 0.60) |
| Gamma (P.1) | 0.51 | (0.44 - 0.61) |
| Delta (B.1.617.2) | 1.03 | (0.95 - 1.11) |
| **Vaccine regimen** |  |  |
| BNT/BNT |  |  |
| Wild-type | 1.00 | (reference) |
| Alpha (B.1.1.7) | 1.01 | (0.78 - 1.31) |
| Beta (B.1.351) | 0.67 | (0.50 - 0.88) |
| Gamma (P.1) | 0.12 | (0.06 - 0.27) |
| Delta (B.1.617.2) | 0.85 | (0.64 - 1.14) |
| ChAd/BNT |  |  |
| Wild-type | 1.00 | (reference) |
| Alpha (B.1.1.7) | 0.74 | (0.66 - 0.83) |
| Beta (B.1.351) | 0.29 | (0.20 - 0.42) |
| Gamma (P.1) | 0.13 | (0.08 - 0.20) |
| Delta (B.1.617.2) | 0.70 | (0.50 - 0.97) |
| ChAd/ChAd |  |  |
| Wild-type | 1.00 | (reference) |
| Alpha (B.1.1.7) | 0.96 | (0.84 - 1.09) |
| Beta (B.1.351) | 0.53 | (0.40 - 0.71) |
| Gamma (P.1) | 0.54 | (0.37 - 0.80) |
| Delta (B.1.617.2) | 0.82 | (0.61 - 1.10) |

Regression output for neutralization against different viral strains. Comparisons are conducted within the separate groups, with neutralization against wild-type within each group as reference. Estimates are multiplicative, i.e. 0.5 corresponds to a 50% reduction in antibody response. Each separate group is a quantile regression model for the median with mixed effect.

## Notes

### Author Declarations

The study was approved by the Swedish Ethical Review Authority (dnr 2020-01653), and written informed consent was obtained from all study participants.

### Summary of Updates

Correction of misprinted numbers in the Abstract ('Findings' section) and a clarification in the legend of table S2

